# Cortical and STN spectral changes during limb movements in PD patients with and without dystonia

**DOI:** 10.1101/2022.01.04.22268757

**Authors:** Joseph W. Olson, Arie Nakhmani, Zachary T. Irwin, Lloyd J. Edwards, Christopher L. Gonzalez, Melissa H. Wade, Sarah D. Black, Mohammad Z. Awad, Daniel J. Kuhman, Christopher P. Hurt, Bart L. Guthrie, Harrison C. Walker

## Abstract

**Background:** Dystonia is an under-studied motor feature of Parkinson disease (PD). Although considerable efforts have focused on brain oscillations related to the cardinal symptoms of PD, whether dystonia is associated with specific electrophysiological features is unclear.

**Objectives:** To investigate subcortical and cortical field potentials at rest and during contralateral hand and foot movements in PD patients with and without dystonia.

**Methods:** We examined the prevalence and distribution of dystonia in PD patients undergoing deep brain stimulation surgery and recorded intracranial electrophysiology from motor cortex and directional electrodes in subthalamic nucleus (STN) both at rest and during self-paced repetitive contralateral hand and foot movements. Wavelet transforms and mixed models characterized changes in spectral content in patients with and without dystonia.

**Results:** Dystonia was highly prevalent at enrollment (61%) and occurred most commonly in the foot. Regardless of dystonia status, cortical recordings display beta (13-30 Hz) desynchronization during movements versus rest, while STN signals show increased power in low frequencies (6.0±3.3 and 4.2±2.9 Hz peak frequencies for hand and foot movements respectively). PD patients with dystonia during DBS surgery displayed greater M1 beta power at rest and STN low frequency power during movements versus those without dystonia.

**Conclusions:** Spectral power in motor cortex and STN field potentials differs markedly during repetitive limb movements, with cortical beta desynchronization and subcortical low frequency synchronization, especially in PD patients with dystonia. Greater knowledge on field potential dynamics in human motor circuits can inform dystonia pathophysiology in PD and guide novel approaches to therapy.

## INTRODUCTION

Dystonia – a twisting, sustained, and often painful involuntary movement – is an under studied motor manifestation of Parkinson disease (PD) [1, 2, 3, 4]. A hallmark of young onset PD, dystonia commonly begins in the foot on the more affected side of the body and spreads to other anatomic locations with disease progression [5, 6, 7, 8]. Accurate estimates of the prevalence of PD-related dystonia have been difficult to obtain. Dystonia may be a presenting symptom of PD in younger patients, occurring in 14-50% in young onset PD patients. PD patients with the parkin mutation have a higher prevalence estimated around 78% [9]. About 30% of levodopa-treated PD patients have some form of dystonia [1], and dystonia severity increases with duration of disease with 2, 12, and 56% prevalence at <5, 6-9, and >10 years duration, respectively [10].

Deep brain stimulation (DBS) of the subthalamic nucleus (STN) is an effective treatment for dystonia related to PD [11, 12, 13], however little is known about its circuit-level electrophysiology. More, however, is known about the electrophysiology of primary dystonia. Growing evidence suggests that primary dystonia is marked by excessive theta and alpha oscillations (<13 Hz) in globus pallidus interna (GPi) at rest, while PD has been associated with excessive power in the beta frequency range (13-30 Hz) [14, 15, 16, 17, 18, 19, 20, 21, 22]. A recent meta-analysis suggests the STN has similar increases in theta and alpha power at rest in primary dystonia compared to PD, with no significant differences in the beta band [23, 24, 25, 26]. Resting local field potentials (LFPs) in primary motor cortex (M1) did not differ significantly for any frequency band in the same meta-analysis [23, 21, 27, 28]. To our knowledge, no studies have compared cortical and subcortical LFPs in PD patients with and without dystonia.

Dystonia is commonly worsened by voluntary movements and can emerge spontaneously in other body parts during voluntary movements. Also, primary dystonia presents in many forms (focal, cervical, generalized, etc.) whereas PD-related dystonia is most common in the foot [29, 30]. Brain electrophysiology during upper and lower limb movements could help characterize motor circuit contributions to PD symptoms such as dystonia. One recent study report that lower and upper limb movement onset coincides with high and low beta desynchronization respectively in both STN and motor cortex [31]. Other studies analyzing spontaneous LFPs during resting, sitting, standing, and walking have found mixed results regarding movement-related STN beta desynchronization [32, 33, 34, 35, 36]. Greater mechanistic knowledge about changes in STN field potentials during specific limb movements could lead to better understanding of pathophysiology and novel strategies for therapy.

Here we investigate LFPs recorded in parallel from sensorimotor cortex and STN during DBS surgery in PD patients with and without dystonia. We recorded from an electrocorticography (ECoG) strip over the ‘hand knob’ of sensorimotor cortex and a directional DBS lead placed at dorsolateral STN. Spectral power at rest and during contralateral hand and foot movements in PD patients, with and without dystonia, were analyzed to test two primary hypotheses: (1) cortical and/or subcortical LFP spectral power differs between hand and foot movements in PD patients (regardless of dystonia status); and (2) cortical and/or subcortical spectral power differs in PD patients with versus without dystonia.

## METHODS

### Recruitment and enrollment

Participants were recruited and studied prospectively as part of the SUNDIAL (SUbthalamic Nucleus DIrectionAL stimulation) study, a randomized, double-blind crossover study contrasting directional versus circular unilateral STN DBS for PD (FDA Investigational Device Exemption G-170063). Dystonia status was not part of the enrollment criteria. Further details on inclusion/exclusion are outlined at https://clinicaltrials.gov/ct2/show/NCT03353688. All subjects provided written informed consent prior to participation with approval from the institutional review board. We included data from all consecutively enrolled participants.

### Dystonia rating

We measured PD motor symptoms with normed, validated clinical instruments, including MDS-UPDRS part 3 and part 4 item 6 (painful ‘off’ dystonia question). There are no comprehensive dystonia rating scales related specifically to PD, therefore the Burke-Fahn-Marsden (BFM) dystonia scale was used to characterize its severity and anatomic distribution [37]. These measures were obtained in a pre-surgery baseline screening visit and longitudinally at 2 and 4 months post-surgery in the practically defined “off” medication state (off dopaminergic medications >12 hours). Additionally, we determined whether dystonia was present or absent intraoperatively based on a neurological examination.

### Electrophysiological recordings

In our practice, DBS surgeries are conducted unilaterally with the patient awake and “off” dopaminergic medications. We recorded LFPs simultaneously from the DBS lead in its final target location in STN and from a linear 6-contact ECoG strip placed temporarily over the hand knob area of ipsilateral sensorimotor cortex (**Figure 1A**) [38]. Strip placement was well tolerated and did not incorporate diuretics or sedative medications. The directional DBS lead (Boston Scientific’s Vercise Cartesia^(tm)^ lead) consists of 8 contacts arranged into 4 rows, each separated by 0.5 mm in a ‘1-3-3-1’ configuration (**Figure 1B**). The lead is placed such that the dorsolateral border of STN lies between the two middle rows of with contacts 2 and 5 facing anteriorly. We acquired surface electromyography (EMG) recordings from contralateral first dorsal interosseous, flexor carpi radialis, and gastrocnemius muscles. All signals were sampled continuously at 25 kHz using a BrainVision actiCHamp amplifier without filters.

**FIGURE 1.**
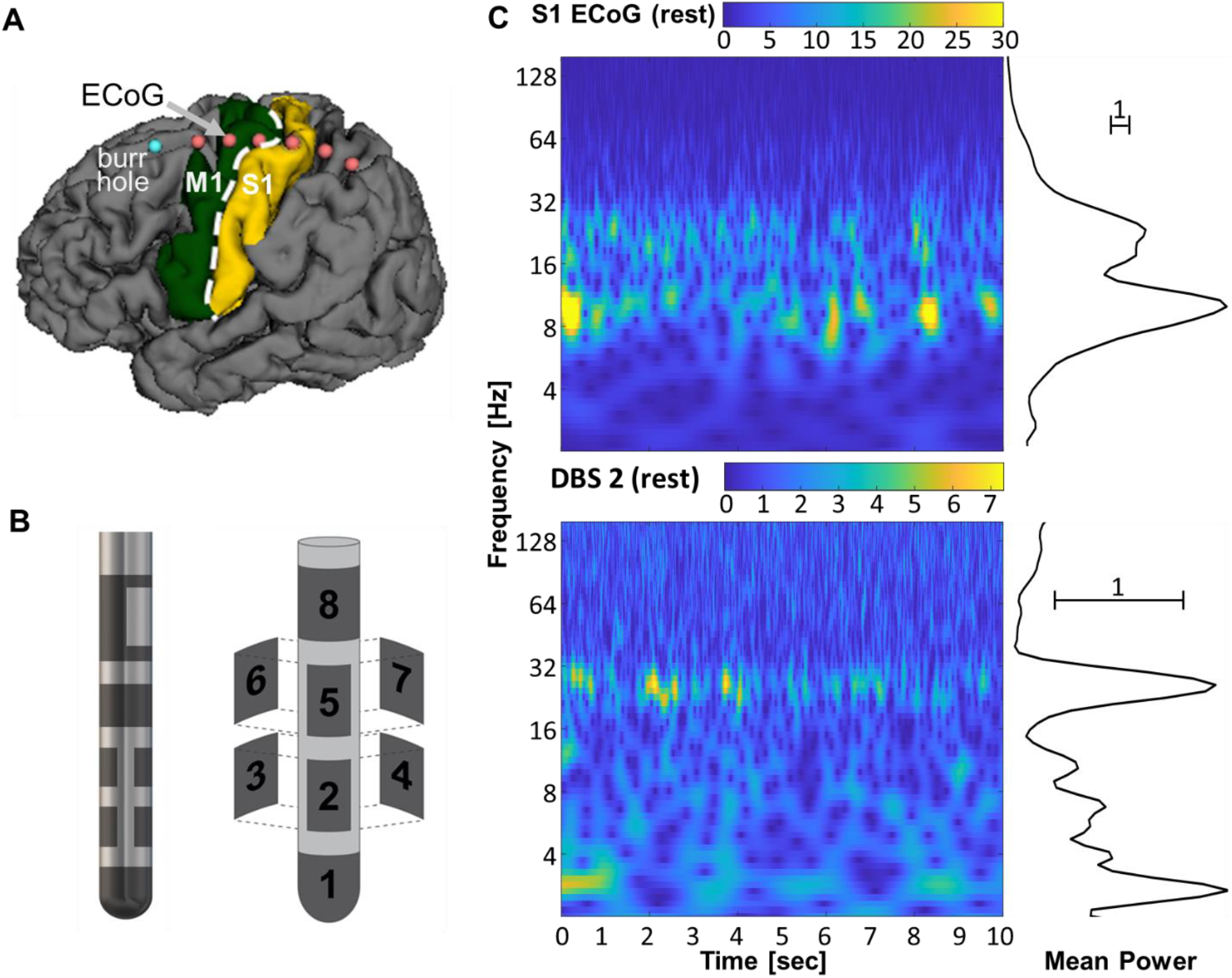
Cortical and subcortical signal recording. **(A)** M1 and S1 regions are overlayed on segmented brain from a pre-op MRI scan. The ECoG strip with 6 contacts is placed over the hand knob area of primary sensorimotor cortex (contacts are outlined in red). It is inserted through the burr hole (blue) during a surgical DBS procedure. **(B)** Directional DBS electrode organization with two rings and two rows of 3 contacts each (1-3-3-1). It is placed along the dorsolateral border of STN as part of routine care. **(C)** Resting state wavelet scalogram example from S1 (above) and DBS contact 2 (below) for a single participant. The mean spectral power estimated on the right is computed by averaging each row of the scalogram over time.

**FIGURE 2.**
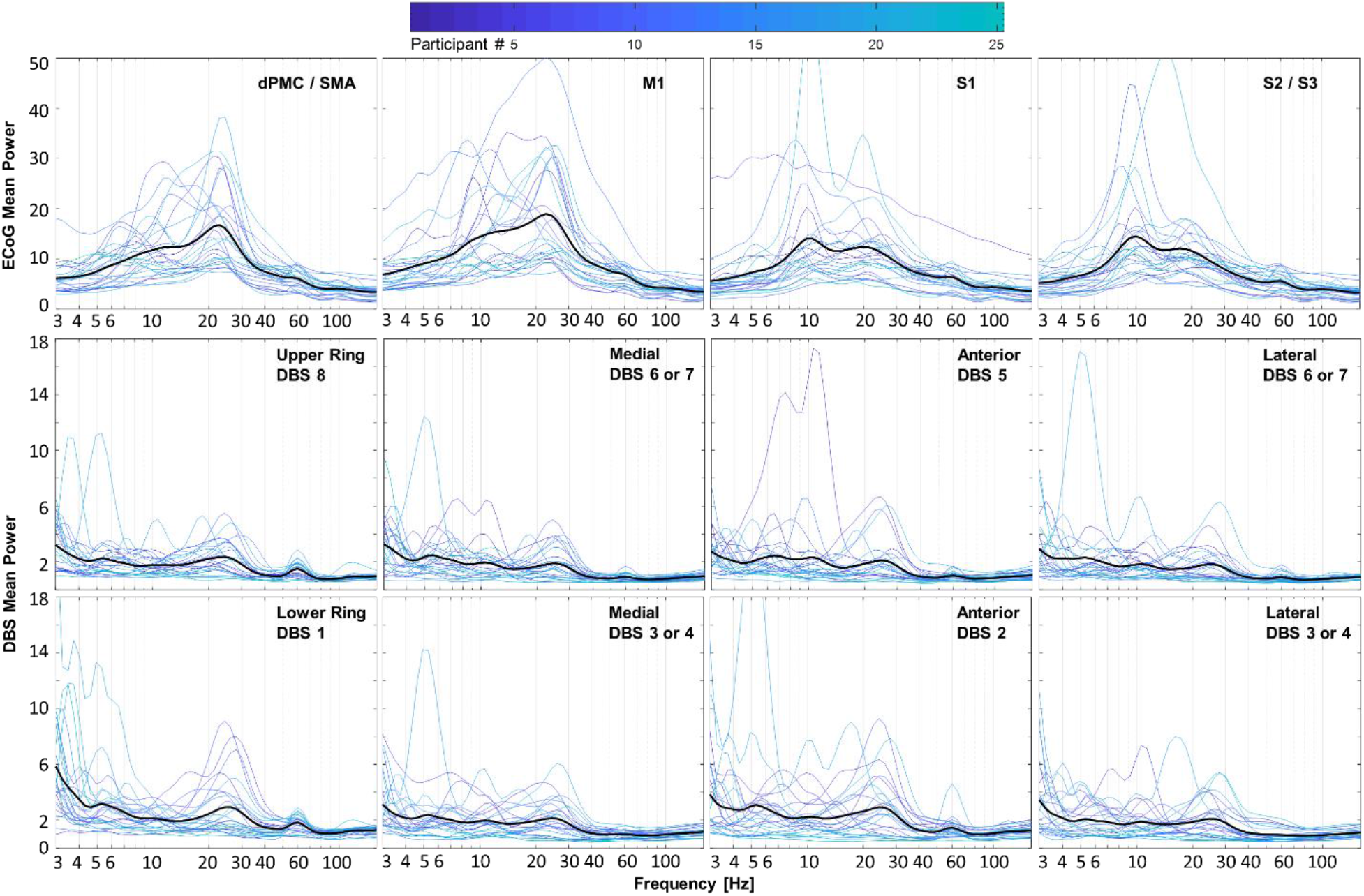
Cortical and subcortical field potentials at rest in PD participants. Spectral power from all electrode contacts over ipsilateral cortex (first row) and the STN region (second and third row) are categorized by anatomical location obtained from imaging, regardless of dystonia status. For each location, there is one plot per participant color-coded by participant number. Means are represented by darker black lines. Beta frequency power is present and relatively large in essentially all electrocorticography contacts, whereas lower beta power is present in most but not all DBS contacts, along with greater relative power at lower frequencies.

### Behavioral testing intraoperative

During intraoperative recording, patients were instructed to rest comfortably over two separate time intervals of one minute each. Verbal instructions for moving the contralateral hand or foot were “open and close your hand as big and fast as possible when you are told ‘go’ until the examiner says ‘stop’,” and “repetitively move your toes and ankle when you are told ‘go’ until the examiner says ‘stop’.” Button press signals corresponded to verbal commands from the examiner (“ready”, “set”, “go”, and “stop”). Hand and foot movements were both recorded in non-consecutive 10 second blocks, with 2 repetitions each. Therefore, the total recording time for rest, foot movement, and hand movement conditions was approximately 2 minutes and 40 seconds (with additional transition time for verbal instructions). Rest and movement intervals were validated post-hoc based upon visualization of EMG recordings.

### Neuroimaging

Pre-op MAGNETOM Prisma MRI scans were segmented with FreeSurfer (Harvard, Martinos Center for Biomedical Imaging) and registered with intra-op O-arm CT for the anatomical localization of ECoG contacts [39]. We used standard methods in Freesurfer to parcellate anatomic regions on the cortical surface. These regions included dorsal premotor cortex (dPMC), supplementary motor area (SMA), primary motor cortex (M1), primary somatosensory cortex (S1), or secondary somatosensory cortex (S2/S3). High-resolution post-op CT was obtained to verify anterior orientation of contacts 2 and 5 on the DBS lead using automated software (Brainlab Inc, Munich, Germany). Directional DBS contacts were classified by row and into medial, anterior, and lateral orientations based on implant hemisphere.

### Digital signal processing and analysis

Signal processing was performed with in-house MATLAB (R2020a; MathWorks, Natick, MA) code and EEGLAB toolbox (UC San Diego, Swartz Center for Computational Neuroscience) [40]. To verify data integrity, we confirmed normal tissue impedance, visualized the raw continuous signals, and generated spectral plots with continuous Morse wavelet transforms (MATLAB function ‘cwt’) on all channels. Invalid signals (with out-of-range impedances) were excluded from subsequent analyses. We separately re-referenced ECoG and DBS signals, using common average reference within each recording modality. All available signals across recording modalities were analyzed, totaling as many as 14 recording sites plus 3 EMG electrodes per participant.

We visualized the LFPs in two ways. First, we generated wavelet scalograms for each contact and then averaged the scalograms over time to obtain spectral magnitude (henceforth called power) across relevant frequencies (**Figure 1C**). To avoid edge artifacts in the wavelet transform computation, we included 2 seconds before and after the movement block, computed the wavelet transform with the expanded signal, and then removed the 4 seconds for padding. Second, we compared the spectra from each DBS and ECoG location at rest and during hand and foot movements with and without dystonia. Frequency ranges for the field potentials are defined as follows: delta (0.5-4 Hz), theta (4-8 Hz), alpha (8-13 Hz), beta (13-30 Hz), gamma (30-70 Hz), and high frequency broadband (hfb) (70-160 Hz). The EMG signal was processed with a 20-1000 Hz band-pass filter and 60 Hz notch filter, followed by rectification and smoothing with a 250 ms Gaussian window. Movement pace was estimated from the EMG by finding peaks (MATLAB function ‘findpeaks’) and using the reciprocal of the average time between peaks.

### Statistical approach

Descriptive statistics including mean, standard deviation, minimum, maximum, and 95% CI were computed for LFP spectral power averaged across all valid contacts within each frequency band for foot, hand, and rest. For each frequency band, we used paired t-tests to compare spectral power during hand and foot movements both to rest and to each other **(Figure 3A)**. To assess spectral differences between patients with or without dystonia **(Figure 5)**, we used linear mixed models [41, 42] with dichotomous predictor for dystonia status, 4th-degree natural polynomials for log-transformed frequency, and the interaction between dystonia status and polynomials as fixed effects to model the wavelet spectrum averages for all recorded channels. Subject-specific intercepts were included as random effects. We identified statistically significant differences between spectrum intervals by identifying overlap within 95% confidence intervals. Linear mixed models were fitted for foot, hand, and rest and CIs were compared based on UPDRS item 4.6, BFM, and surgical exam. No statistical corrections for multiple comparisons were performed.

**FIGURE 3.**
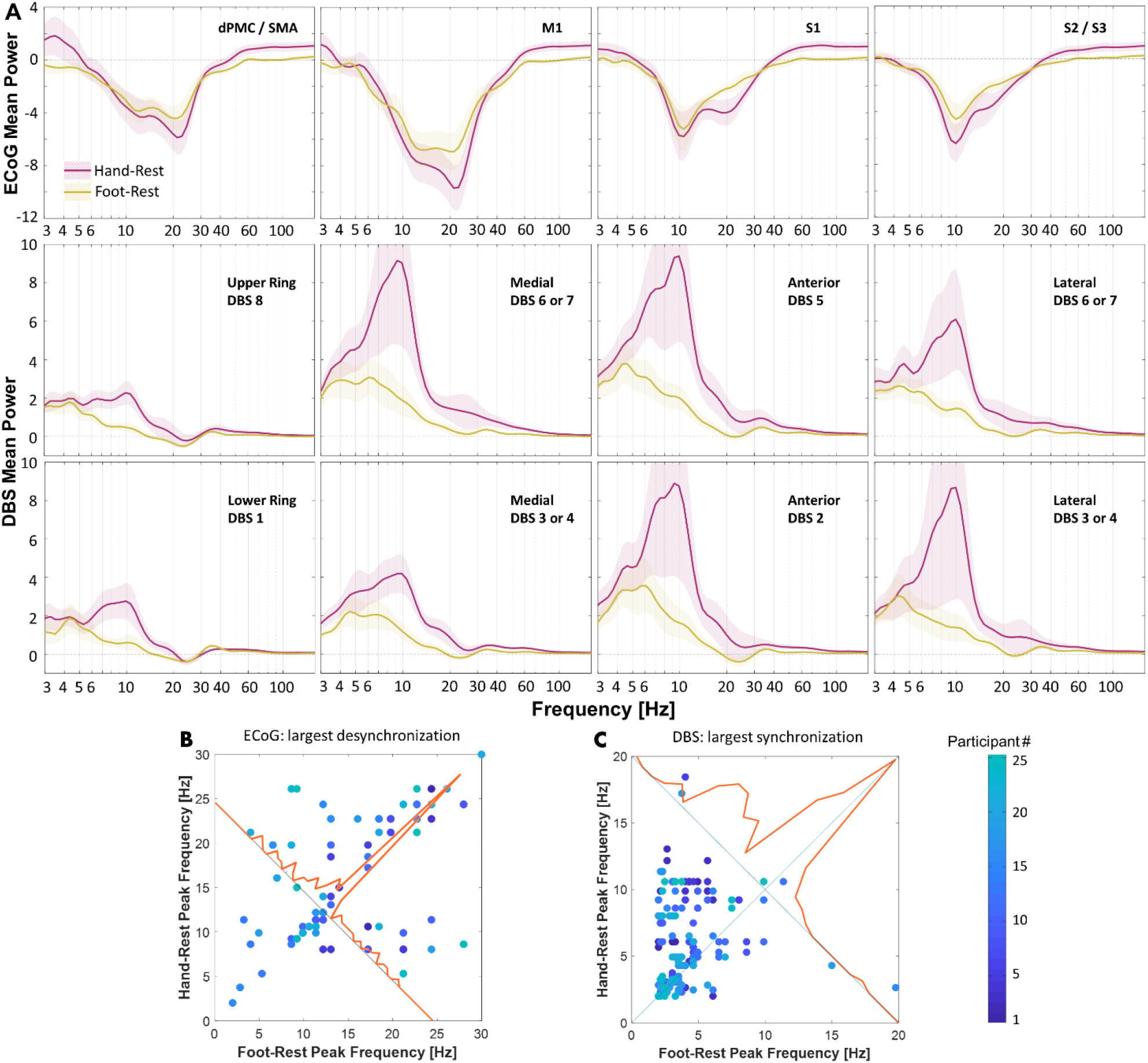
Cortical and subcortical spectral perturbations in PD participants during continuous, repetitive hand and foot movements minus rest. **(A)** Spectral power for all participants (n=25) and recording sites are summarized in row 1 for sensorimotor cortex and rows 2 and 3 for dorsal and ventral DBS contacts (mean across participants ± standard error). Red and yellow traces correspond to hand and foot signals, respectively. **(B, C)** Frequencies that displayed the strongest deviations during movement versus rest in cortex and STN (i.e., either desynchronization or synchronization). If the spectral perturbations are identical, observations will appear on or near the unity line (diagonal line). **(B)** Frequencies corresponding to power minima in dPMC/SMA, M1, S1, S2/S3 contacts during foot versus hand movements. **(C)** Frequencies corresponding to power maxima from all DBS contacts during foot versus hand movements.

**FIGURE 4.**
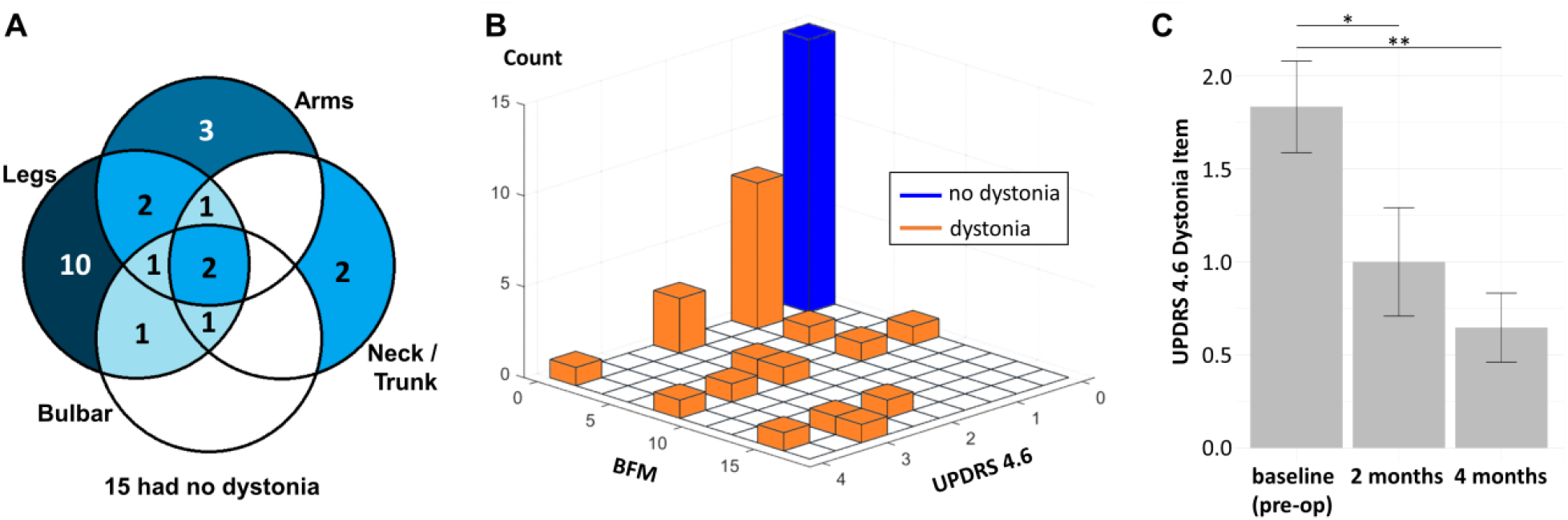
Prevalence and anatomic distribution of dystonia in PD participants evaluated for DBS surgery during baseline visit, and clinical outcomes. **(A)** Venn diagram of anatomic distribution of dystonia clustered into four groups (arms, legs, bulbar, neck/trunk). The number in each circle intersection denotes the number of participants who had the corresponding parts affected by the dystonia (self-reported or discovered by Burke-Fahn-Marsden dystonia rating scale). Fifteen out of 38 participants had no dystonia at baseline. **(B)** Three-dimensional histogram of UPDRS dystonia item 4.6 (painful dystonia scale) versus BFM total score. Orange bars denote participants with dystonia and the blue bar denotes participants with no dystonia. **(C)** Dystonia motor outcomes following DBS. Error bars indicate standard error. Both 2 months and 4 months are significant from baseline (p=0.002; p<0.001).

**FIGURE 5.**
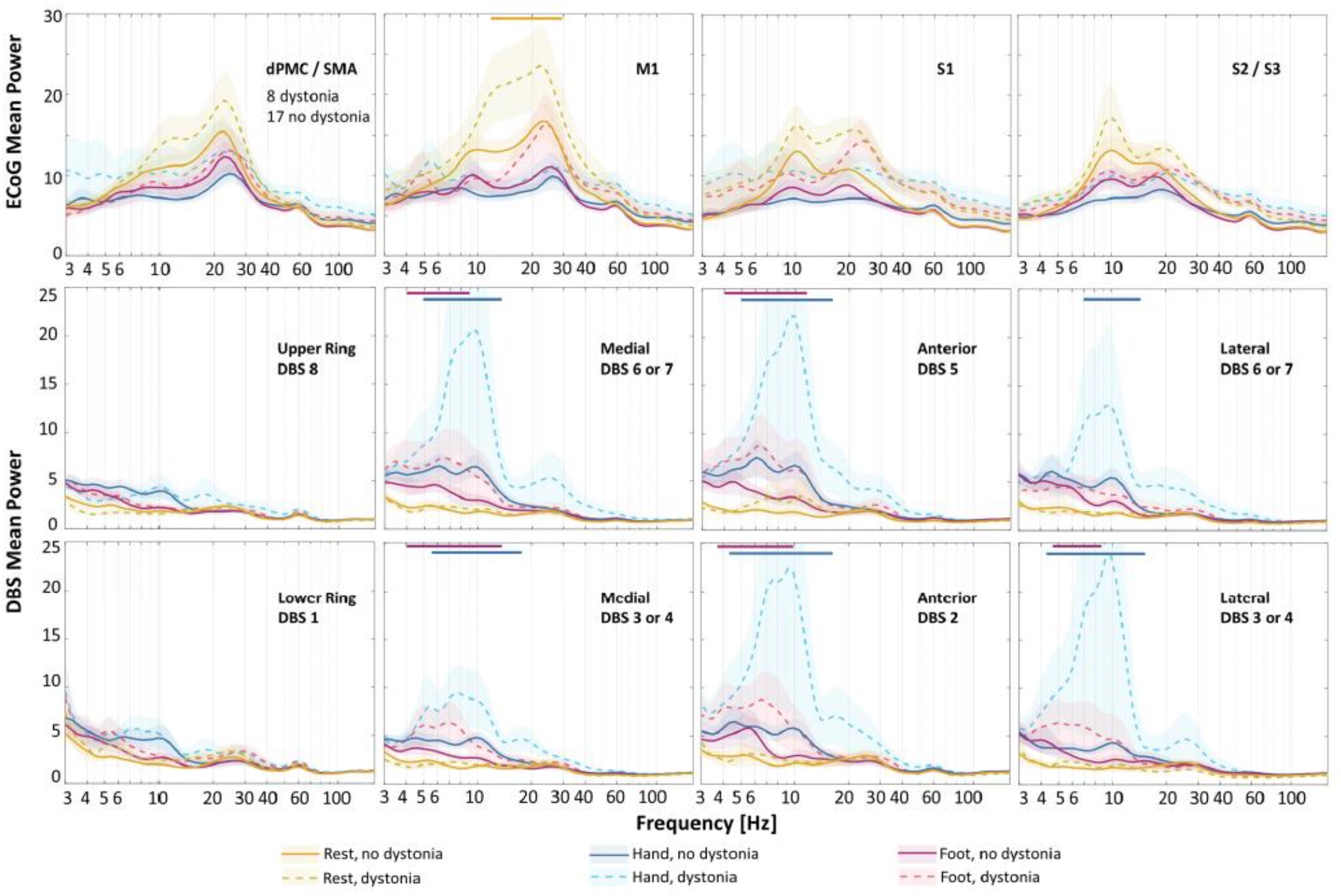
Cortical and subcortical spectral perturbations during rest, hand, and foot movements in PD participants with versus without dystonia during DBS surgery. Spectral power from ECoG contacts in ipsilateral cortical (first row) and DBS contacts in STN region (second row) are categorized by presence/absence of dystonia during DBS surgery (mean ± standard error). Dashed and solid lines represent average spectral power in PD patients with and without dystonia, respectively. Yellow, blue, and red colors correspond to rest, hand, and foot movements. Shaded standard errors are for visual purposes only and do not necessarily reflect the statistical confidence intervals of non-normal variance. Horizontal colored bars above the plots correspond to frequency intervals with statistically significant differences.

### Data Sharing

All data are available on the Data Archive for the Brain Initiative (DABI) at https://dabi.loni.usc.edu/home.

## RESULTS

### Demographics

We studied intraoperative LFPs in 28 of 38 (74%) participants who met entry criteria for the SUNDIAL trial. Exclusions related to the following: ≤30% improvement UPDRS part 3 “on” versus “off” PD medications (*n*=4); Beck Depression Inventory >10 (*n*=1); multidisciplinary committee recommended globus pallidus rather than STN target (*n*=2); medical condition requiring future MRI (*n*=1); health insurance refused payment for surgery with an investigational device (*n*=1); and voluntary withdrawal (*n*=1). Of the 28 participants with recorded LFPs, 3 had corrupted or missing data, yielding 25 participants for subsequent analyses.

### Spectral perturbations from rest during hand and foot movements

The EMG recordings of all 25 participants were obtained to verify their resting intervals. Spectral analyses of resting field potentials revealed peaks in the beta frequency range (13-30 Hz) in both sensorimotor cortex and STN, with approximately five times the magnitude at cortical recording sites (**Figure 2**). To isolate changes in spectral power related to movement, we subtracted resting spectra from movement spectra within each channel (**Figure 3A**). Cortical LFPs demonstrate broadband power changes during repetitive voluntary hand and foot movements compared to rest. Alpha and beta power of cortical LFPs decrease markedly during both movements, while hfb power increases during hand movements only (Foot. theta: -1.0 ±1.9, p=0.014; alpha: -4.0 ± 3.8, p<0.001; beta: -3.4 ± 2.3, p<0.001; gamma: -0.6 ± 0.9, p=0.004. Hand. alpha: -4.8 ± 5.0, p<0.001; beta: -4.6 ± 3.6, p<0.001; hfb: 1.0 ± 1.7, p=0.006. Mean ± standard deviation, t-test). Spectral power differs significantly between foot and hand movements in the beta, gamma, and high frequency bands (beta: 1.2 ± 2.2, p=0.014; gamma: -0.6 ± 1.2, p=0.028; hfb: -0.9 ± 1.1, p<0.001).

Interestingly, spectral power in STN signals during repetitive limb movements displayed an almost opposite behavior. Theta, alpha, and low beta spectral power increased during movements compared to rest, while the higher beta power increased slightly or did not change (Foot. delta: 2.0 ± 1.7, p<0.001; theta: 2.3 ± 2.9, p<0.001; alpha: 1.3 ± 2.3, p=0.010; gamma: 0.2 ± 0.2, p<0.001; hfb: 0.1 ± 0.0, p<0.001. Hand. delta: 2.6 ± 2.3, p<0.001; theta: 4.1 ± 5.8, p=0.002; alpha: 5.5 ± 11.8, p=0.029; gamma: 0.4 ± 0.8, p=0.012; hfb: 0.1 ± 0.2, p<0.001). Furthermore, spectral power during foot and hand movements differed from each other in theta, alpha, and hfb frequency ranges (theta: -1.7 ± 3.6, p=0.023; alpha: -4.2 ± 9.8, p=0.043; hfb: -0.1 ± 0.2, p=0.023).

In addition to measuring spectral power, we also characterized which peak frequencies displayed the most prominent changes during limb movements. We measured frequencies with the maximal desynchronization during hand and foot movements versus rest in the ECoG signals (**Figure 3B**). Given that the STN field potentials were inverted and showed increases in spectral power, we instead measured frequencies with maximal synchronizations (**Figure 3C**). In most cases, maximal cortical desynchronization occurred at similar frequencies during foot versus hand movements (14.4 ± 6.6 Hz and 15.8 ± 6.7 Hz, respectively, mean ± standard deviation, p=0.460, paired t-test). In contrast, maximal synchronization in the STN region occurred at lower peak frequencies during foot versus hand movements (4.2 ± 2.9 Hz and 6.0 ± 3.3 Hz respectively, p=0.046). We found no statistically significant correlation between LFP frequency and movement rate during hand or foot movements (**supplementary Figure 1**).

### Dystonia prevalence and clinical features

Dystonia status was assessed based on participant report (UPDRS item 4.6), the BFM score at a motor screening visit, and neurological examination during DBS surgery. We provide all available behavioral data on PD-related dystonia, even in participants who were screen failures for the intraoperative component of the study to provide a more comprehensive accounting of dystonia phenomenology in patients with Parkinson’s disease. In total, dystonia was present based on history, examination, or both in 23 of 38 participants (61%). Dystonia occurred most, but not exclusively, in the foot (18/23, 78%) (**Figure 4A**). Among survey responses from baseline screening, 8 of 17 participants (47%) report worsening of dystonia during various movements, whereas other reported dystonia primarily at rest (**supplementary Table 1**). During surgery, 8 of 25 participants (32%) had subjective or visible dystonia “off” meds. Although UPDRS item 4.6 correlates linearly with BFM total score (F = 8.337, *p* = 0.006, adjusted R-square = 0.114) (**Figure 4B**), of the 22 participants with dystonia based on UPDRS 4.6, only 12 (45%) had dystonia “off” medications at their screening visit based on BFM, illustrating the transient nature of dystonia in PD. Interim analyses provide evidence that unilateral STN DBS improves dystonia symptoms, based on changes in UPDRS item 4.6 at 2- and 4-months post-op versus pre-op baseline (F= 24.127, p<0.001, linear mixed model with dependent variable UPDRS 4.6 and fixed effect time, subject-specific random intercept) (**Figure 4C**). UPDRS part 3 total score ‘off’ meds show clinically significant motor improvement of 33.7, 32.6, and 37.5% at 2, 4, and 6 months versus preoperative baseline (baseline score 49.2 ± 2.38 reduced by16.6±1.6, 16.0 ± 1.6, and 18.6± 1.6, p< 0.001, respectively, linear mixed effects model) [43].

### Electrophysiological biomarkers of dystonia in PD

All patients were classified as having or not having dystonia based on neurological exam during surgery. Wavelet spectrum averages for each channel were plotted at rest and during hand, and foot movements (**Figure 5**). There was a greater M1 beta power (13-29 Hz) at rest in PD patients with versus without dystonia (95% confidence interval). There were no other significant differences in spectral power between PD patients with and without dystonia at other cortical regions or frequency range across conditions, at our level of statistical power. We observed no significant differences in STN power at rest in PD patients with versus without dystonia. During both foot and hand movements, we detected greater STN low frequency power (∼4-12 Hz and ∼5-15 Hz, respectively) in PD patients with dystonia in directional DBS contacts only (95% confidence intervals). We also compared spectral power of PD patients with versus without dystonia using UPDRS 4.6 and BFM rating scales (**supplementary Figure 2**). Group differences were greatest based on the presence/absence of dystonia at the time of the recordings during surgery.

## DISCUSSION

Here we report three primary findings, each related primarily to activity in the theta and alpha frequency ranges. First, we use intraoperative LFPs to provide evidence that PD patients with versus without dystonia display greater low frequency STN power during limb movements and greater beta power in M1 at rest (**Figure 5**). Second, regardless of dystonia status, foot movements increased STN spectral power predominantly in the theta range, whereas contralateral hand movements yielded greater synchrony at alpha frequencies (**Figure 3**). Finally, sensorimotor cortex and basal ganglia field potentials differ markedly during repetitive, naturalistic voluntary movements, with sustained beta desynchronization in cortex and broadband activation at low frequencies in the STN (**Figure 3**).

### Dystonia pathophysiology in subcortical and cortical field potentials

Although we are unaware of prior studies on this topic in PD patients, studies on primary dystonia suggest excessive theta and alpha power in basal ganglia at rest compared to PD (regardless of their dystonia status). We did not identify similar differences in our sample at rest but did observe excessive STN theta and alpha power during limb movements (**Figure 5**). Dystonia symptoms often worsen with voluntary movements, such dystonia pathophysiology might be more pronounced or easily detected in STN during movements (**supplementary Table 1**). We also found increased M1 beta power at rest in PD patients with versus without dystonia, where other groups have used TMS to identify abnormal short-term plasticity in patients with both primary dystonia and PD [44]. Of note, we found increases in high frequency broadband power during hand movements only, which confirms placement of the electrocorticography strip over the hand area of motor cortex [45, 46] (**Figure 3**).

### Low frequency STN activity during upper and lower limb movements

In the STN, repetitive limb movements yielded broadband increases in low frequency power, with foot and hand movements manifesting primarily in the theta and alpha frequency ranges, respectively (**Figure 3**). While this differs somewhat from a study analyzing LFPs time-locked to movement onset [31], others have implicated low frequency activity in STN during lower limb movements, as well [32, 34]. While some argue that these low frequency signals in basal ganglia represent movement artifacts [34], we and others [32] contend that they likely arise from brain signals for several reasons. First, ECoG and DBS signals in our paradigm show opposite changes in spectral power during movements versus rest (**Figures 3 and 5**), yet the wires that transmit the signals are bundled together and are sampled by the same amplifier. Second, we observed no correlations between peak frequencies in the LFPs and the pace or rate of movements across participants (**supplementary Figure 1**).

### Beta desynchronization is greater in sensorimotor cortex than in STN

Consistent with prior studies, we found prominent beta band frequencies at rest in cortex, along with large and sustained beta desynchronizations during voluntary movements [47]. Resting beta power was also observed in STN, although it was considerably smaller in power and present less consistently (**Figure 2**). In agreement with a similar recent study, we did not observe beta desynchronization during repetitive movements (**Figure 3**) [48]. These results may differ from prior studies reporting beta desynchronization in STN at movement onset because our paradigm studied naturalistic, self-paced repetitive movements rather than cued single movements. Repetitive or complex movements can display beta-rebound phenomena [49, 50], and ‘microlesion’ effects from acute lead placement might alter the local responses, as well [51]. Regardless, beta desynchronization in cortex was more robust and sustained during sustained repetitive movements versus in STN.

### Clinical translational significance

Dystonia is an underrecognized symptom of PD, with historical prevalence estimates at approximately 30% [1]. Although we observed dystonia during surgery in 31% of our sample, other available measures indicate a total prevalence of at least 61%. Another group reported higher dystonia prevalence (71%) in PD patients undergoing DBS surgery, as well [11]. Underestimates could arise from the fluctuating nature of dystonia, lack of data capture in routine assessments, and/or lack of recognition of dystonia symptoms. Another contributor is likely sampling bias, such that PD patients with dystonia are more likely to receive DBS than those without. Given its painful nature and relatively high prevalence in this group of patients, additional electrophysiology studies on PD-related dystonia are warranted. Simultaneous cortical and directional subcortical field potential recordings in this study provide new pathophysiological knowledge on spectral perturbations during repetitive limb movements in PD patients with and without dystonia. Additionally, multimodal recordings from this study could inform development of emerging adaptive DBS technologies, given that primary motor cortex and basal ganglia are leading candidate regions for closed-loop sensing [52].

### Study Limitations

Our study has some potential limitations. Time and physical constraints during surgery sometimes limit data collection, and acute implant of the DBS lead introduces microlesion effects that could alter the electrophysiology [51]. Therefore, elements of these studies should be repeated with implanted devices with sensing capabilities in the chronic setting. Dystonia was most commonly present in the foot, yet our ECoG placements were over the hand knob of motor cortex. While we did see increased beta power at rest in dystonia patients in M1, this nevertheless might have limited our view of cortical physiology related to dystonia. We used EMG recordings to characterize movement states, but future studies would likely benefit from video recordings to better capture time-locked movement dynamics. Although our sample is considerably larger than most prior efforts in the field, this is an initial implementation of our analytic methods with these broadband signals. As such, no corrections for multiple comparisons were employed. Thus, our results should be interpreted with some caution.

## CONCLUSIONS

Spectral perturbations in primary motor cortex and STN field potentials differ substantially during repetitive arm and leg movements, with prominent beta desynchronization in cortex and increased theta and alpha power in STN, especially in PD patients with dystonia. Greater knowledge on field potential dynamics in human motor circuits promise to inform dystonia pathophysiology in PD and guide novel approaches to therapy.

## Data Availability

All data is available on the Data Archive for the Brain Initiative (DABI) at https://dabi.loni.usc.edu/home.

## ACKNOWLEDGMENT

We would like to thank Matthew J. Nelson for scientific discussions and advice.

## AUTHORS’ ROLES

a. Experimental design
b. Data collection
c. Data analysis and/or making figures
d. Writing of manuscript
e. Edits of manuscript

|  |  |  |
| --- | --- | --- |
| 454 | Joseph W. Olson | b, c, d, e |
| 455 | Arie Nakhmani | a, c, d, e |
| 456 | Zachary T. Irwin | a, b, e |
| 457 | Lloyd J. Edwards | c, d, e |
| 458 | Christopher L. Gonzalez | b, c, e |
| 459 | Melissa H. Wade | b, e |
| 460 | Sarah D. Black | c, e |
| 461 | Mohammad Z. Awad | b, e |
| 462 | Daniel J. Kuhman | a, b, e |
| 463 | Christopher P. Hurt | a, b, e |
| 464 | Bart L. Guthrie | b, e |
| 465 | Harrison C. Walker | a, b, c, d, e |

## FINCANCIAL DISCLOSURES (PAST 12 MONTHS)

**466 FINANCIAL DISCLOSURES (PAST 12 MONTHS)**
|  |  |  |
| --- | --- | --- |
| 467 | Stock Ownership in medically-related fields: | No authors |
| 468 | Intellectual Property Rights: | No authors |
| 469 | Consultancies: | No authors |
| 470 | Expert Testimony: | No authors |
| 471 | Advisory Boards: | No authors |
| 472 | Employment: | UAB (all authors) |
| 473 | Partnerships: | No authors |
| 474 | Inventions: | No authors |
| 475 | Contracts: | No authors |
| 476 | Honoraria: | No authors |
| 477 | Royalties: | No authors |
| 478 | Patents: | No authors |
| 479 | Grants: | NIH, MJFF |
| 480 | Other: | No authors |

---

**SUPPLEMENTARY TABLE 1.** Patient experiences with dystonia surveyed pre-surgery.

| **General description of dystonia** | **Activities that worsen dystonia** | **Body part affected by dystonia** |
| --- | --- | --- |
| 'All the time when meds wear off.' | 'No specific activities.' | 'Both feet. Toes curl.' |
| 'Unclear to participant. It seems random.' | 'Standing, walking, resting.' | 'Left, middle, fingers, and toes on right foot. Right fingers will draw.' |
| 'Leg morning and evening (morning before medicines work).' | 'Ocular is brought on by bright lights. Leg activity is brought on if he thinks about it' | 'Leg and eye' |
| 'Every 4 hours when meds wear off' | 'She experiences it at rest.' | 'Right toes. Right leg. Both wrists. Her neck.' |
| 'Shortly after dose of medicine until it starts wearing off.' | 'Sitting, at rest' | 'Left foot' |
| 'Only notice it when sitting on couch watching TV at the end of the day' | 'Only notice it when resting' | 'Left and right feet areas affected' |
| 'When tired' | 'Walking will bring it on' | 'Left foot' |
| 'Does not happen at specific time of day' | 'Sitting and rest' | 'Both legs' |
| '1st thing in the morning when meds are off.' | 'At rest, also hands will activate if reading or holding something; Foot at rest, with meds off.' | 'Both feet, both hands/wrists and neck.' |
| 'Afternoon' | 'At rest' | 'Neck' |
| 'When medicines are off' | 'Anything that requires continuous effort/force' | 'Right side hand and toes' |
| 'When medicines wear off' | 'Rest' | 'Neck, trunk, right arm and right foot' |
| 'All the time.' | 'Always.' | 'Left toes and neck.' |
| 'Starts after participant takes the first dosage of medicine' | 'At rest' | 'Left leg' |
| 'Night, afternoon' | 'Lack of sleep, lack of exercise.' | 'Right shoulder, hip, foot' |
| 'Occurs often' | 'Walking, anxiety' | 'Right foot' |
| 'Does not occur often' | 'Rest' | 'Left arm' |

**SUPPLEMENTARY FIGURE 1.**
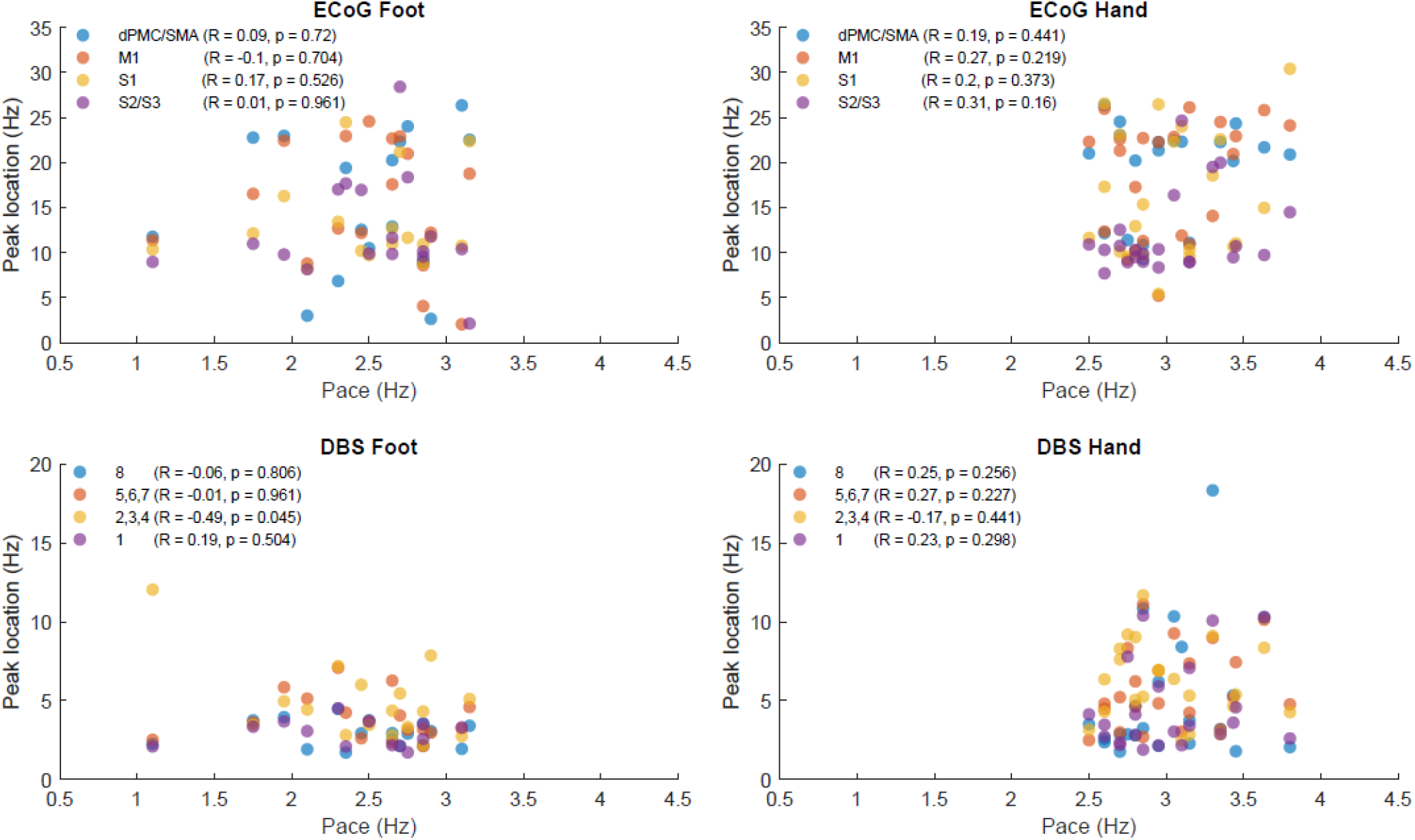
Correlation between the movement pace and spectral extrema. Above are frequencies of maximum cortical desynchronization during movement compared to rest (FIGURE 3). For each cortical region, Pearson correlation is given with p-values not adjusted for multiple comparisons. Below are frequencies of maximum subcortical synchronization during movement compared to rest.

**SUPPLEMENTARY FIGURE 2.**
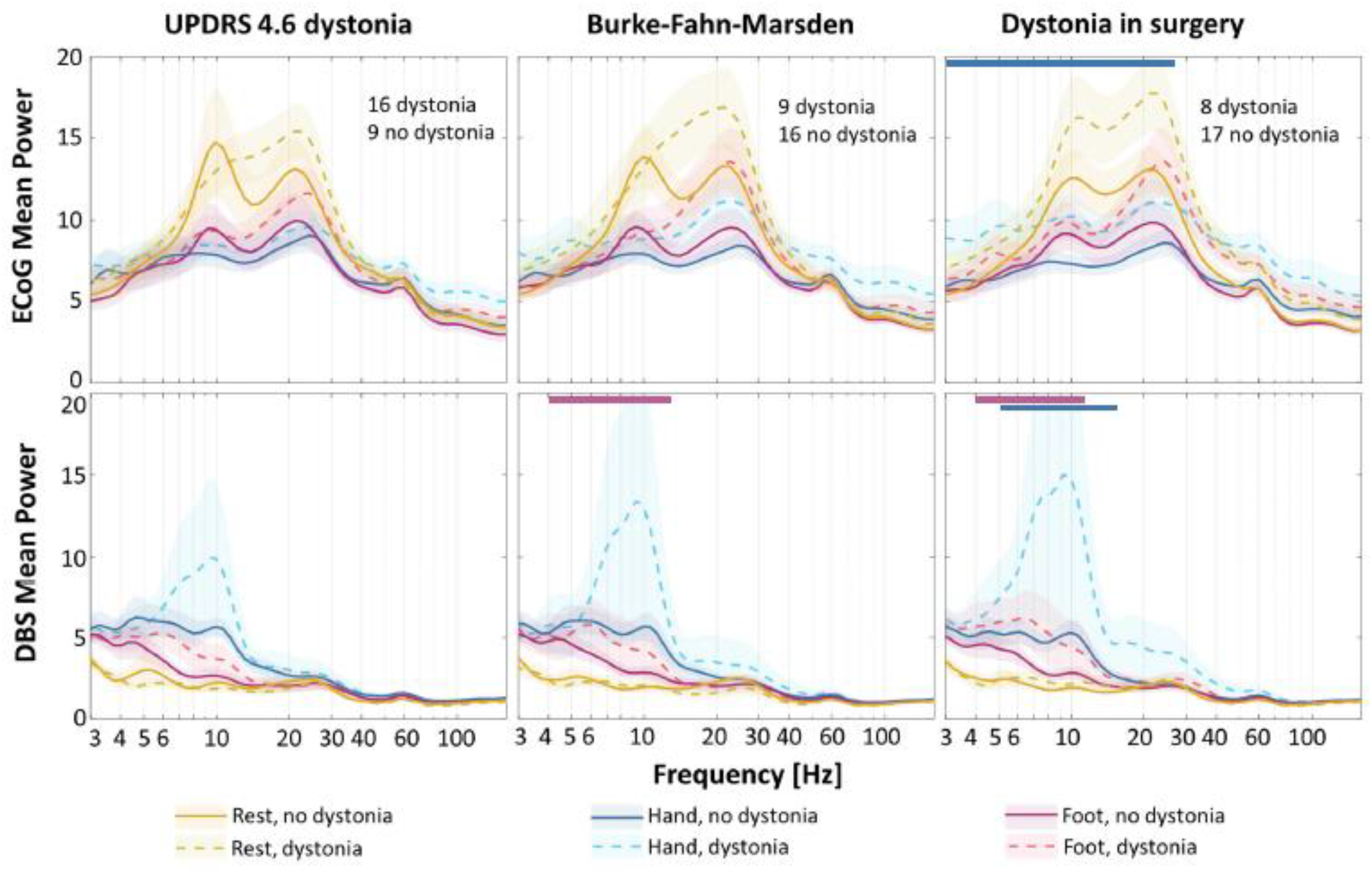
Averaged cortical and subcortical spectral perturbations during rest and limb movements in PD participants with versus without dystonia determined by different dystonia rating scales. Spectral power from all electrode contacts over ipsilateral sensorimotor cortex (first row) and STN region (second row) are categorized by presence/absence of dystonia historically (UPDRS 4.6), at pre-operative baseline assessment (BFM), and during DBS surgery (mean ± standard error). Dashed and solid lines represent average spectral power in PD patients with and without dystonia, respectively. Yellow, blue, and red colors correspond to rest, hand, and foot movements. Shaded standard errors are for visual purposes only and do not necessarily reflect the statistical confidence intervals of non-normal variance. Horizontal colored bars above the plots correspond to frequency intervals with statistically significant differences. During foot movement, we observed statistically significant differences (p<0.05) between STN low frequency power in PD patients with vs without dystonia when using BFM (4-12 Hz) and surgical exam (4-11 Hz) but not UPDRS. Significant differences (p<0.05) in STN power were also observed during hand movement when using surgical exam classification (5-15 Hz). No significant differences in STN power were observed during rest for any dystonia classification. Furthermore, there was a significant difference (p<0.05) in cortical spectral power during hand movement using surgical exam classification (3-25 Hz). Otherwise, there were no observed differences in cortical spectral power between PD patients with and without dystonia.

